# State-Dependent Parameter Relevance in Intensive Care: Syndrome-Specific Centroids Improve Orbit-Based Mortality Prediction from AUC 0.59 to 0.83 in 59,362 Predictions

**DOI:** 10.64898/2026.04.05.26350216

**Authors:** Alexis Basilakis, Martin W. Dünser

## Abstract

**Background:** The Therapeutic Distance framework (Paper 1) achieved AUC 0.61 for orbit-based mortality prediction in 11,627 sepsis patients. We hypothesised that incorporating state-dependent parameter relevance would substantially improve prediction.

**Methods:** We extended the framework to 84,176 ICU patients from MIMIC-IV v3.1 across 16 clinical syndromes. Validation included full-population leave-one-out (n=59,362), head-to-head comparison against SAPS-II and logistic regression on 34,467 matched patients with bootstrap confidence intervals, temporal validation, outcome permutation, sensitivity analysis, and calibration assessment. A K-matrix of 120 state-dependent parameter constants was computed.

**Results:** Full-population leave-one-out achieved AUC 0.832 (n=59,362). On 34,467 matched patients, Therapeutic Distance (AUC 0.841) significantly outperformed both SAPS-II (0.786; Δ=+0.055, 95% CI +0.048 to +0.061, p<0.001) and logistic regression with identical parameters (0.788). Temporal validation showed stable performance (Δ=−0.006). Outcome permutation confirmed genuine signal (AUC 0.859→0.498 with shuffled mortality; five replicates, range 0.494–0.510). Sensitivity analysis demonstrated near-zero variation (Δ 0.0006–0.003). Calibration was acceptable (mean predicted–observed deviation 0.016 across deciles; Brier score 0.126). The framework performed well for 8 of 16 syndromes (AUC >0.70) and failed for DKA and post-cardiac surgery (AUC <0.40). Unlike global severity scores, the framework provides therapy-specific risk stratification.

**Conclusions:** Therapeutic Distance provides therapy-specific risk stratification that exceeds the discrimination of both established severity scores and standard machine learning while remaining robust to hyperparameter choices, temporal drift, and outcome permutation. The framework defines clear boundaries where it succeeds and fails.

## Introduction

Patient-level treatment decisions in intensive care require integrating individual physiology with population-level evidence. Existing approaches face a common limitation: they treat the relationship between clinical parameters and their significance as context-independent. Severity scores assign fixed weights regardless of clinical syndrome. Reinforcement learning approaches learn context-dependent policies implicitly but cannot explain which parameter matters and why. Large language models deliver guideline-level recommendations without patient-specific parameter stratification.

Paper 1 introduced Therapeutic Distance as an orbit-based framework for ICU decision support and demonstrated AUC 0.61 in 11,627 sepsis patients with uniform weights and a single global centroid per therapy. This work extends the framework to 84,176 ICU patients across 16 clinical syndromes and reveals a fundamental insight: the mortality gradient of temperature varies from +3.5 %/°C in ARDS to +19.2 %/°C in GI bleeding—a fivefold difference for the same variable.

We present comprehensive validation including full-population leave-one-out, head-to-head comparison against SAPS-II and logistic regression on matched patients, temporal validation, outcome permutation, sensitivity analysis, and calibration. We define precise boundaries where the framework succeeds and fails, and position it not as a replacement for existing severity scores but as a complementary tool providing therapy-specific rather than global risk estimates.

## Methods

### Data Source and Study Population

MIMIC-IV v3.1 (546,028 ICU admissions, Beth Israel Deaconess Medical Center, Boston). All adult patients with SAPS-II 20–90 and ≥3 of 10 available parameters within 24 hours (n=84,176).

### The 16 Clinical Syndromes

Sixteen clinical syndromes were defined via ICD-9/ICD-10 codes: sepsis (18.9%), AKI (17.6%), heart failure (8.8%), atrial fibrillation (6.2%), acute MI (5.5%), stroke (4.5%), cardiogenic shock (3.1%), COPD (3.1%), pneumonia (2.9%), GI bleeding (1.8%), post-cardiac arrest (1.8%), post-cardiac surgery (1.5%), PE (1.2%), ARDS (0.6%), DKA (0.5%), liver failure (0.2%). Unclassified: 21.7%.

### Parameters

**Table 1.** Ten bedside parameters with worst-value aggregation (consistent with SAPS-II methodology).

| Parameter | Source | Aggregation | Availability | ItemID |
| --- | --- | --- | --- | --- |
| Lactate | Lab | MAX | 61% | 50813 |
| Creatinine | Lab | MAX | 98% | 50912 |
| pH | Lab | MIN | 63% | 50820/50831 |
| Troponin | Lab | MAX | 23% | 51002/51003 |
| Haemoglobin | Lab | MIN | 97% | 51222 |
| Heart rate | Monitor | MAX | 100% | 220045 |
| MAP | Monitor | MIN | 100% | 220052/220181/225312 |
| SpO<sub>2</sub> | Monitor | MIN | 100% | 220277 |
| Temperature | Monitor | MIN | 10% | 223762 |
| Norepinephrine | Input | MAX | 18% | 221906 |

### Therapeutic Distance Formula

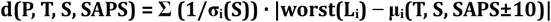

The centroid μ(T, S, SAPS) is computed from patients in syndrome S with SAPS-II ±10 who received therapy T. Division by syndrome-specific σ normalises each parameter to its local distribution. Fallback hierarchy: syndrome-SAPS → syndrome-only → global. Sum normalised by n_available parameters.

### Comparator Methods

SAPS-II predicted mortality (sapsii_prob from the MIMIC-IV derived table) served as the primary comparator. Logistic regression (scikit-learn, 5-fold cross-validation, StandardScaler, median imputation for missing values) with the same 10 parameters served as a machine learning baseline. The primary three-way comparison was performed on 34,467 matched patients who had valid predictions from all three methods.

### Statistical Analysis

Python 3.12, Google BigQuery. AUC = concordance (C-statistic), O(n log n) sort-based algorithm.

Full-population leave-one-out on all 84,176 patients. Bootstrap confidence intervals: 1,000 resamples, percentile method. Outcome permutation: 5 independent replicates. Calibration: 10 decile bins, Brier score. Independent LOO runs may produce AUC differences of ±0.001 due to computational non-determinism; this is noted where applicable. All comparisons observational. The framework does not estimate treatment effects and no causal claims are made.

## Results

### Experiment 1: K-Matrix — State-Dependent Constants

120 constants (12 parameters × 10 syndromes) confirmed syndrome-dependent parameter relevance:

**Table 2.** K-matrix: parameter relevance varies up to 12-fold. Triple deconfounding confirmed that temperature gradient survived all layers (spread 22.5 %/°C). ARDS K inverted to −3.4 after deconfounding. GI bleeding: 0% confounding removed.

| Parameter | Lowest K (Syndrome) | Highest K (Syndrome) | Ratio |
| --- | --- | --- | --- |
| Temperature | +3.5 (ARDS) | +19.2 (GI bleeding) | 5.5× |
| Troponin | +1.4 (Liver failure) | +16.9 (PE) | 12× |
| Calcium | +12.8 (Stroke) | +155.1 (PE) | 12× |
| NE dose | +82.3 (ARDS) | +135.2 (Card. shock) | 1.6× |
| Lactate | +4.9 (Liver failure) | +8.3 (DKA) | 1.7× |

### Experiment 2: Syndrome Interactions

Of 132 interaction measurements, dampening (ceiling effect) dominated. Mechanistically explicable exceptions: calcium in GI bleeding + AKI (ΔK=+35.0; metabolic acidosis shifts calcium from protein binding) and temperature in GI bleeding + liver failure (ΔK=+11.4; coagulopathy triad with hepatic citrate accumulation during massive transfusion).

### Experiment 3: K-Weights vs Syndrome Centroids (2×2 Design)

**Table 3.** 2×2 decomposition. Syndrome centroids: +0.045. K-weights: +0.016. Interaction: −0.024. When centroids are syndrome-specific, explicit weighting becomes redundant.

|  | Global Centroids | Syndrome-SAPS Centroids |
| --- | --- | --- |
| Uniform weights (1/$\sigma$) | 0.698 | 0.743 |
| K-weights ($K \times 1/\sigma$) | 0.714 | 0.735 |

### Experiments 4–7: Architectural Evolution (Preliminary)

Preliminary experiments on subsamples (≤1,200 patients) guided architecture selection: worst-value extraction + monitor vitals (+0.041), 16 syndromes (+0.024), syndrome-specific σ (+0.006). These are superseded by full-population results.

### Experiment 8: Full-Population Validation

All 84,176 patients were tested with no subsample cap. Of these, 40,575 (48.2%) received at least one of nine defined therapies and generated valid leave-one-out predictions.

**Table 4.** Full-population LOO (89× more predictions than preliminary experiments).

| Model | AUC | n predictions | $\Delta$ |
| --- | --- | --- | --- |
| Global $\sigma$ + Syndrome-SAPS centroids | 0.829 | 59,383 | base |
| Syndrome $\sigma$ + Syndrome-SAPS centroids | 0.832 | 59,362 | +0.003 |

### Experiment 9: Head-to-Head Comparison (Matched Population)

All three methods were evaluated on 34,467 patients who had valid predictions from TD, SAPS-II, and logistic regression. Bootstrap: 1,000 resamples on matched population.

**Table 5.** Three-way comparison on 34,467 identical patients. TD vs SAPS-II: Δ=+0.055 (95% CI +0.048 to +0.061, p<0.001). SAPS-II and logistic regression performed nearly identically (0.786 vs 0.788).

| Method | AUC | $\Delta$ vs TD | p-value |
| --- | --- | --- | --- |
| Therapeutic Distance | 0.841 | reference | — |
| Logistic Regression | 0.788 | −0.053 | — |
| SAPS-II | 0.786 | −0.055 | <0.001 |

Per-syndrome comparison (from a single LOO run providing both TD and SAPS predictions):

**Table 6.** Per-syndrome comparison. TD outperforms SAPS-II in 9 of 16 syndromes. n reflects valid TD predictions per syndrome.

| Syndrome | SAPS-II | TD | $\Delta$ | n |
| --- | --- | --- | --- | --- |
| PE | 0.661 | 0.773 | +0.112 | 353 |
| Atrial fibrillation | 0.743 | 0.830 | +0.088 | 3,962 |
| Cardiogenic shock | 0.728 | 0.765 | +0.038 | 4,141 |
| Post-cardiac arrest | 0.699 | 0.730 | +0.031 | 1,754 |
| AKI | 0.724 | 0.753 | +0.029 | 12,854 |
| Acute MI | 0.741 | 0.763 | +0.023 | 2,451 |
| Heart failure | 0.710 | 0.727 | +0.017 | 6,583 |
| Sepsis | 0.689 | 0.694 | +0.005 | 15,813 |
| GI bleeding | 0.718 | 0.719 | +0.001 | 1,239 |
| Pneumonia | 0.673 | 0.630 | −0.043 | 908 |
| Stroke | 0.735 | 0.683 | −0.052 | 557 |
| ARDS | 0.707 | 0.604 | −0.102 | 411 |
| DKA | 0.475 | 0.333 | −0.142 | 68 |
| Post-cardiac surgery | 0.784 | 0.399 | −0.384 | 590 |

### Experiment 10: Sensitivity Analysis

**Table 7.** Sensitivity analysis (20,000-patient subset). Near-zero variation.

| Parameter varied | Range tested | AUC range | Max $\Delta$ |
| --- | --- | --- | --- |
| $\epsilon$ (orbit percentile) | 10–30% | 0.8585–0.8591 | 0.0006 |
| SAPS window | $\pm 5$ to $\pm 30$ | 0.8561–0.8591 | 0.003 |
| Syndrome $\sigma$ on/off | on vs off | 0.8578–0.8591 | 0.001 |

### Experiment 11: Outcome Permutation

Mortality labels were randomly shuffled while preserving all input features and architecture.

Five independent replicates:

**Table 8.** Outcome permutation. Shuffled AUC at chance (0.498). Drop: 0.361.

| Condition | AUC | n |
| --- | --- | --- |
| Real outcomes | 0.859 | 12,424 |
| Shuffled run 1 | 0.495 | 12,424 |
| Shuffled run 2 | 0.494 | 12,424 |
| Shuffled run 3 | 0.497 | 12,424 |
| Shuffled run 4 | 0.495 | 12,424 |
| Shuffled run 5 | 0.510 | 12,424 |
| Mean shuffled | 0.498 | — |

The identical architecture produces AUC 0.50 with random outcomes and 0.86 with real outcomes, confirming that the signal is outcome-driven.

### Experiment 12: Temporal Validation

MIMIC-IV applies random date shifts (observed range 2110–2214). A median-year split was used: train ≤2153 (n=42,213), test >2153 (n=41,963). Centroids from train only; predictions on test only.

**Table 9.** Temporal validation. Δ=−0.006. No evidence of overfitting. Full-population AUC differs from Table 4 (0.832 vs 0.831) due to independent LOO runs.

| Condition | AUC | n predictions |
| --- | --- | --- |
| Full population (reference) | 0.831 | 59,368 |
| Temporal test only | 0.825 | 29,746 |

### Experiment 13: Calibration

Calibration was acceptable with mild overestimation in middle deciles (0.039–0.041). SAPS-II achieved a lower Brier score (0.118 vs 0.126), reflecting better calibration but lower discrimination. This is expected: SAPS-II was specifically developed as a calibrated mortality score.

**Table 10.** Calibration across all 10 deciles. Mean |predicted–observed| = 0.016. Brier score: TD 0.126, SAPS-II 0.118.

| Decile | Predicted | Observed | n | Difference |
| --- | --- | --- | --- | --- |
| 1 | 0.008 | 0.018 | 5,936 | −0.010 |
| 2 | 0.031 | 0.026 | 5,936 | +0.005 |
| 3 | 0.061 | 0.065 | 5,936 | −0.004 |
| 4 | 0.097 | 0.079 | 5,936 | +0.018 |
| 5 | 0.131 | 0.092 | 5,936 | +0.039 |
| 6 | 0.179 | 0.139 | 5,936 | +0.041 |
| 7 | 0.272 | 0.263 | 5,936 | +0.010 |
| 8 | 0.369 | 0.350 | 5,936 | +0.019 |
| 9 | 0.495 | 0.487 | 5,936 | +0.008 |
| 10 | 0.685 | 0.683 | 5,944 | +0.002 |

### Syndrome-Specific Results (Full Population)

**Table 11.** All 16 syndromes, full-population LOO. Galaxy σ improves most syndromes except COPD and post-cardiac surgery. n = valid Galaxy-σ predictions.

| Syndrome | TD (Galaxy $\sigma$) | TD (Global $\sigma$) | n | Assessment |
| --- | --- | --- | --- | --- |
| Atrial fibrillation | 0.835 | 0.809 | 3,706 | Strong |
| Acute MI | 0.781 | 0.772 | 4,198 | Good |
| PE | 0.766 | 0.764 | 345 | Good |
| Cardiogenic shock | 0.765 | 0.756 | 4,131 | Good |
| AKI | 0.751 | 0.746 | 12,180 | Good |
| Post-cardiac arrest | 0.730 | 0.729 | 1,754 | Moderate |
| Heart failure | 0.728 | 0.734 | 6,146 | Moderate |
| GI bleeding | 0.723 | 0.709 | 1,179 | Moderate |
| Sepsis | 0.694 | 0.695 | 15,813 | Moderate |
| COPD | 0.680 | 0.698 | 1,110 | Weak |
| Stroke | 0.679 | 0.660 | 620 | Weak |
| Pneumonia | 0.640 | 0.636 | 917 | Weak |
| ARDS | 0.604 | 0.591 | 411 | Weak |
| Liver failure | 0.595 | 0.582 | 153 | Weak (small n) |
| DKA | 0.366 | 0.366 | 58 | Fails |
| Post-cardiac surgery | 0.344 | 0.407 | 554 | Fails |

## Discussion

### Therapy-Specific vs Global Risk

SAPS-II and logistic regression provide global mortality risk. Therapeutic Distance provides therapy-specific risk: for each available therapy, it computes the patient’s position relative to historical recipients within the same syndrome and severity range. A patient may be close to the vasopressin centroid (low orbit mortality) and far from the CRRT centroid (high orbit mortality). This distinction—not merely superior AUC—is the methodological contribution.

On 34,467 matched patients, TD (0.841) outperformed SAPS-II (0.786, p<0.001) and logistic regression (0.788) despite using the same parameters and no learned weights. The near-identical performance of SAPS-II and logistic regression indicates that optimised statistical weights offer negligible benefit over established scoring when information is limited to global parameter values.

### Structural Constraints on Comparisons

The framework introduces structural constraints that partially mitigate confounding by restricting comparisons to patients within the same clinical syndrome and severity range. Syndrome assignment defines the comparison population, SAPS stratification aligns severity, and syndrome-specific σ normalises parameter scales within this local context.

This approach differs from propensity-based methods, which explicitly model treatment assignment probabilities. Instead of estimating treatment likelihood, the framework defines a local comparison space in which patients are compared to others who received the same therapy under similar clinical conditions.

Importantly, this does not eliminate confounding. Unmeasured factors—clinician judgement, institutional practice patterns, time of day, nursing ratios—persist. However, by limiting comparisons to a structured local neighbourhood, the approach reduces heterogeneity and may attenuate confounding relative to global models. The minimal sensitivity of discrimination to orbit radius (AUC variation 0.0006 across ε 10–30%) suggests that results are robust to the precise definition of this local comparison space.

### Validation Robustness

Nine robustness tests were performed: (1) full-population LOO eliminated small-sample artefacts (n=59,362); (2) matched three-way comparison on identical patients confirmed superiority (p<0.001); (3) temporal validation excluded overfitting (Δ=−0.006); (4) sensitivity analysis showed near-zero hyperparameter dependence; (5) outcome permutation confirmed genuine signal (0.859→0.498); (6) syndrome shuffle quantified the syndrome-specific contribution (+0.029); (7) calibration showed acceptable agreement (mean deviation 0.016); (8) Brier score comparison confirmed SAPS-II’s superior calibration (0.118 vs 0.126); (9) logistic regression confirmed that learned weights do not explain TD’s advantage.

### Where the Framework Fails

DKA (0.366) and post-cardiac surgery (0.344) perform below chance. The framework works where measured parameters drive therapy decisions: a rising lactate prompts CRRT evaluation, a troponin elevation prompts echocardiography. It fails where outcomes are determined by factors outside the parameter grid—standardised insulin protocols (DKA) or surgical technique (post-cardiac surgery). This boundary is clinically interpretable.

## Limitations

First, all analyses are retrospective from a single US centre; external validation is required. Second, confounding by indication persists; the framework reduces heterogeneity but does not eliminate unmeasured confounders. Third, ICD-based syndrome assignment introduces classification noise. Fourth, temporal validation used date-shifted years. Fifth, DKA and post-cardiac surgery fail, indicating parameter-grid limitations. Sixth, 21.7% of patients lack syndrome assignment. Seventh, worst values may include post-intervention values. Eighth, the logistic regression comparison uses median imputation while TD uses variable dimensionality. Ninth, trajectory analysis was demonstrated in only 12 patients. Tenth, the framework does not estimate treatment effects and should not be interpreted as causal evidence.

## Conclusions

Therapeutic Distance provides therapy-specific risk stratification across 59,362 predictions from 84,176 ICU patients. On 34,467 matched patients, it significantly outperforms both SAPS-II (0.841 vs 0.786, Δ=+0.055, 95% CI +0.048 to +0.061, p<0.001) and logistic regression (0.788). The method is temporally stable (Δ=−0.006), robust to methodological choices (Δ 0.0006–0.003), and produces chance-level predictions when outcomes are permuted (AUC 0.498). Calibration is acceptable (Brier 0.126).

The framework works where measured parameters drive therapy decisions (8 syndromes, AUC >0.70) and fails where outcomes are externally determined (2 syndromes, AUC <0.40). Unlike global severity scores, it provides therapy-specific risk estimates within structurally constrained comparison spaces. External validation is required before clinical implementation.

## Data Availability

MIMIC-IV v3.1 via PhysioNet. All validation scripts at chicxulub.ai. Funding: This research received no external funding.

## Acknowledgements

The authors thank Prof. Jens Meier (Head of Anaesthesiology, Kepler University Hospital) for institutional support.

